# Predicting COVID-19 peaks around the world

**DOI:** 10.1101/2020.04.24.20078154

**Authors:** Constantino Tsallis, Ugur Tirnakli

## Abstract

The official data for the time evolution of active cases of COVID-19 pandemics around the world are available online. For all countries, a peak has been either observed (China and South Korea) or is expected in near future. The approximate dates and heights of those peaks imply in important epidemiological issues. Inspired by similar complex behaviour of volumes of transactions of stocks at NYSE and NASDAQ, we propose a *q*-statistical functional form which appears to describe satisfactorily the available data of all countries. Consistently, predictions become possible of the dates and heights of those peaks in severely affected countries unless efficient treatments or vaccines, or sensible modifications of the adopted epidemiological strategies, emerge.

---

It is possible to predict the thermostatistical properties of uncountable physical systems at thermal equilibrium through the one-body distribution 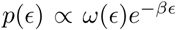, where *ω*(*ε*) is the density of states as a function of the energy ε, multiplied by the celebrated Boltzmann factor *e*^−^*^βε^*, *β* being the inverse temperature. The function *ω*(*ε*) comes from mechanical considerations (classical, quantum, relativistic) related with the number of degrees of freedom, and does not depend on the temperature; the exponential weight comes instead from standard statistical-mechanical considerations. In many cases it is, either exactly or approximatively, 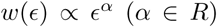. For thermostatistical properties of the stationary- or quasi-stationary-state of wide classes of complex systems, the Boltzmann factor is to be generalised into the *q*-exponential factor 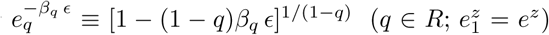 [1–3]. This procedure yielded quite satisfactory results for the high-frequency stock-market in NYSE, NASDAQ and elsewhere [4–6].

Let us focus now on the data available for the COVID-19 pandemics. Briefly after the beginning of the pandemics, several studies analysing the available data, and employing different models and candidate functions, started to appear in the literature [7–12]. Most of them are interested in the behaviour of total cases and fatality curves. We will concentrate here on the analysis of the active cases and deaths *per day*. The inspection of the public data [13] (updated on a daily basis), in particular of the time evolution of the number *N* of active cases (surely a lower bound of the unknown actual numbers) showed a rather intriguing similarity with the distributions of volumes of stocks. Along this line we adopt the following functional form for each country or region:

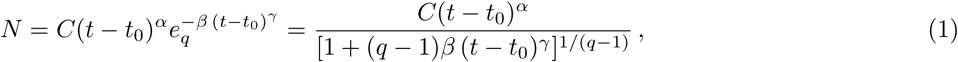

with *C* > 0; *α* > 0; *β* > 0; *γ* > 0, *q* > 1 and *t*_0_ ≥ 0. The constant *t*_0_ indicates the first day of appearance of the epidemics in that particular region; it is conventionally chosen to be zero for China; for the other countries it is the number of days elapsed between the appearance of the first case in China and the first case in that country. The normalising constant *C* reflects the total population of that particular country. For *α* = 0, if *γ* = 1 we recover the standard *q*-exponential expression; if *γ* = 2, it is currently referred in the literature as *q*-Gaussian; for other values of *γ*, it is referred to as stretched *q*-exponential. Through the inspection of the roles played by the four nontrivial parameters, namely (*α*, *β*, *γ*, *q*), it became rather transparent that (*α*, *β*) depend strongly on the epidemiological strategy implemented in that region, in addition to the biological behaviour of the coronavirus in that geographical climate. In contrast, the parameters (*γ*, *q*) appear to be sort of universal, mainly depending on the coronavirus. Therefore, we investigate several countries, which do not reach their peaks yet, with the basic assumption that these two parameter would not change much from one country to another and we fixed these values at the values that we determine for China since this country has already had nearly the full evolution of the pandemics. This assumption seems to be working. For other contries whose peaks are already reached, we use the same functional form (1) but adjusting all parameters for a better fitting. The results of China and South Korea are given in Fig. 1. It is evident that, although the functional form (1) does yield satisfactory results for both China and South Korea, the (*γ*, *q*) parameter values differ somewhat for each of these countries. On the other hand, as can be easily seen from Fig. 2, our assumption is corroborated by several countries that we have numerically analysed. In Table 1 we present the forecasted dates and heights of the peaks, as well as the values of the fitting parameters using the data accumulated until 28 of April, 2020.

**FIG. 1:**
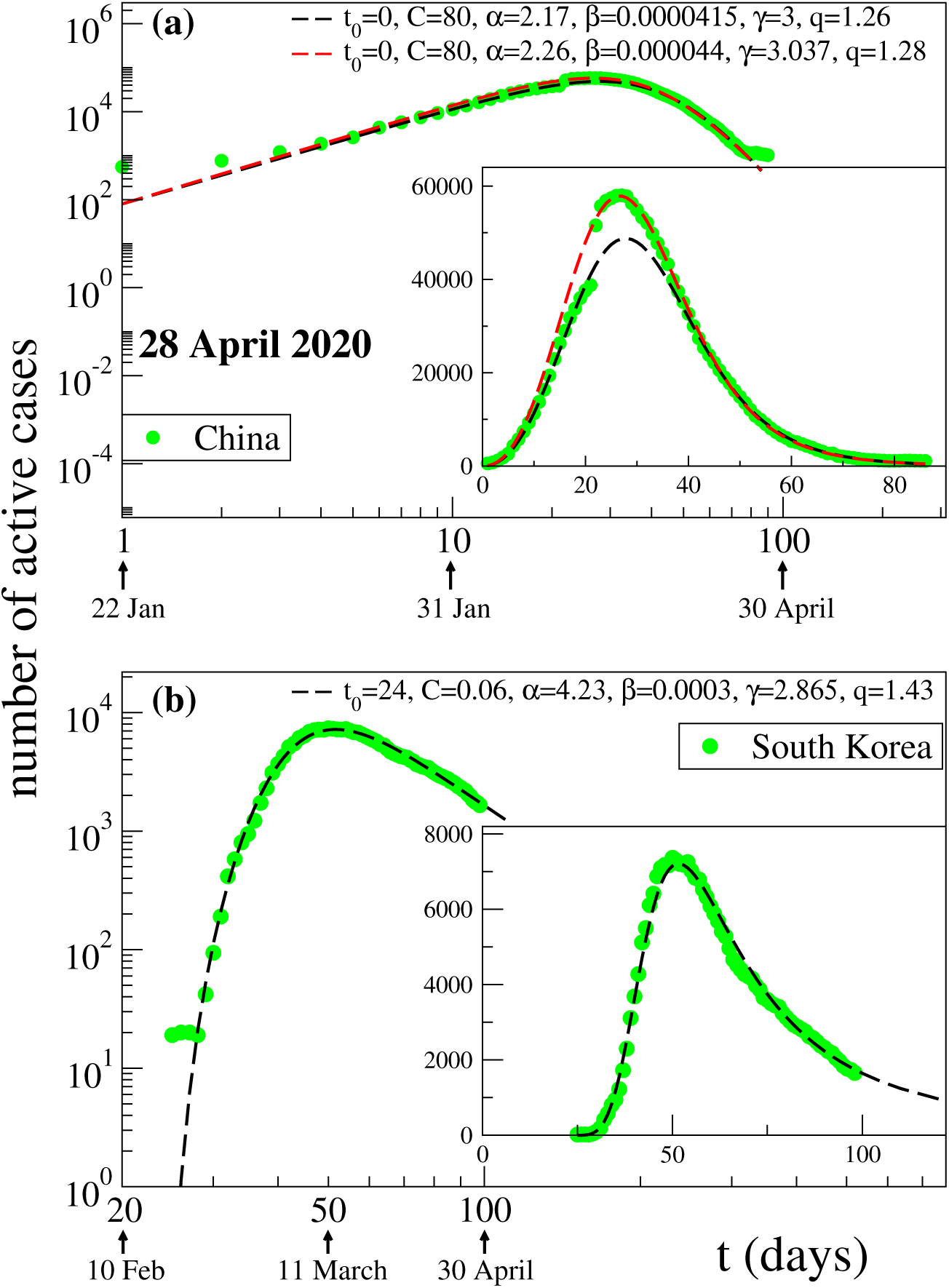
Two possible fits for the COVID-19 active cases evolution in (a) China and (b) South Korea: the dots are the available data at 28 April 2020, and the lines are fittings using Eq. (1). For China a strange kink is present in the ascending part of the data curves, which makes that it is not possible to make a single fitting which would satisfactorily account for both increasing and decreasing parts. We present here two log-log representation of different curves which describe either the increasing part or the decreasing part but not both. Inset: linear-linear representation of the same data.

**FIG. 2:**
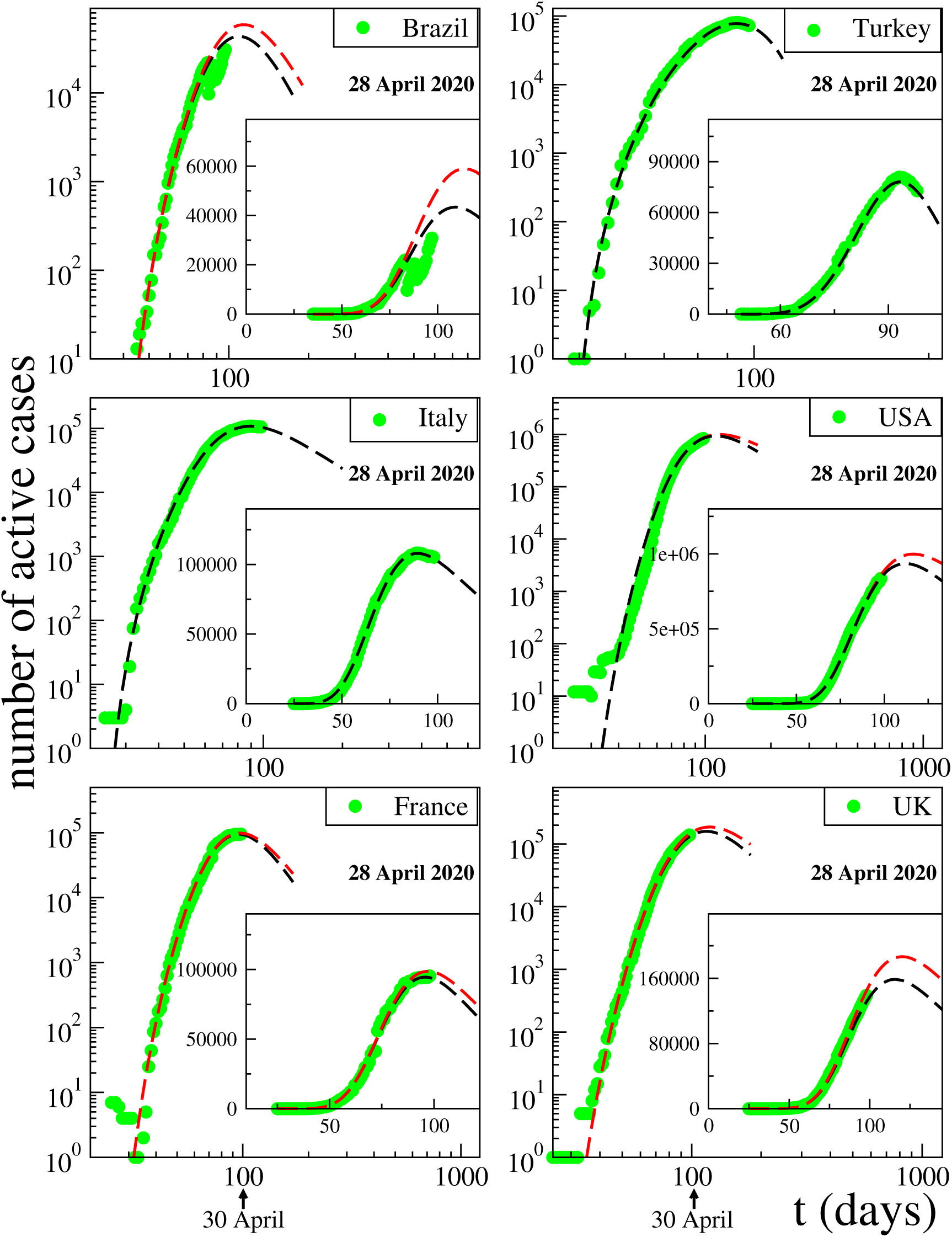
Fittings of the active cases data available in 28 April 2020 for various severely a ected countries around the world with Eq. (1). The fitting parameters *γ* and *q* are fixed at the values of China for all countries which do not reach their peak values yet. Notice that, in the case of Brazil, a disruption occurred in the publicly available data in middle April. We do not know the cause of this. Coincidentally, however, precisely in those days the President of Brazil decided to change his Minister for Health.

**TABLE I:** The values of the parameters and predictions for the maximum number of active cases and for the day at which the maximum will be achieved. The second line in the column of prediction interval of active cases peak height shows the same data divided by the total population of the country.

| $\gamma$ and $q$ parameters are fixed at China's values if peak is not achieved yet | | | | | | | | |
| --- | --- | --- | --- | --- | --- | --- | --- | --- |
| Country | $t_0$ | $C$ | $\alpha$ | $\beta$ | $\gamma$ | $q$ | Prediction interval<br>of peak height | Prediction interval<br>of peak day |
| Brazil | 34 | $1.55 \times 10^{-5}$ | 5.65 | $8.00 \times 10^{-6}$<br>$8.00 \times 10^{-6}$ | 3<br>3.037 | 1.26<br>1.28 | [43400, 58965]<br>[0.00021, 0.00028] | [109, 114]<br>[09 May, 14 May] |
| Turkey | 48 | $8.0 \times 10^{-3}$ | 4.625 | $1.52 \times 10^{-5}$ | 3.025 | 1.02 | achieved at<br>80808 | achieved on day 93<br>23 April |
| Italy | 24 | $4.0 \times 10^{-3}$ | 4.68 | $1.10 \times 10^{-5}$ | 3.083 | 1.43 | achieved at<br>106100 | achieved on day 96<br>26 April |
| USA | 24 | $3.4 \times 10^{-9}$ | 8.60 | $1.65 \times 10^{-5}$<br>$1.46 \times 10^{-5}$ | 3<br>3.037 | 1.26<br>1.28 | [934680, 997830]<br>[0.0028, 0.0030] | [112, 117]<br>[12 May, 17 May] |
| France | 24 | $1.6 \times 10^{-6}$ | 6.56 | $9.20 \times 10^{-6}$<br>$1.19 \times 10^{-5}$ | 3<br>3.037 | 1.26<br>1.28 | [94551, 98726]<br>[0.0014, 0.0015] | [96, 97]<br>[26 April, 27 April] |
| UK | 24 | $1.2 \times 10^{-8}$ | 7.60 | $9.60 \times 10^{-6}$<br>$8.00 \times 10^{-6}$ | 3<br>3.037 | 1.26<br>1.28 | [158700, 186610]<br>[0.0024, 0.0028] | [116, 120]<br>[16 May, 20 May] |

We also test our formula (1) and assumption for the evolution of deaths per day. Again we fixed the (*γ*, *q*) parameter values at China’s results, which can be seen in Fig. 3, and tried to fit the data of the same 6 countries. It is quite surprising to see that the results given in Fig. 4 seem to suggest that the evolution of death per day is also possible to be fitted without changing these two parameters.

**FIG. 3:**
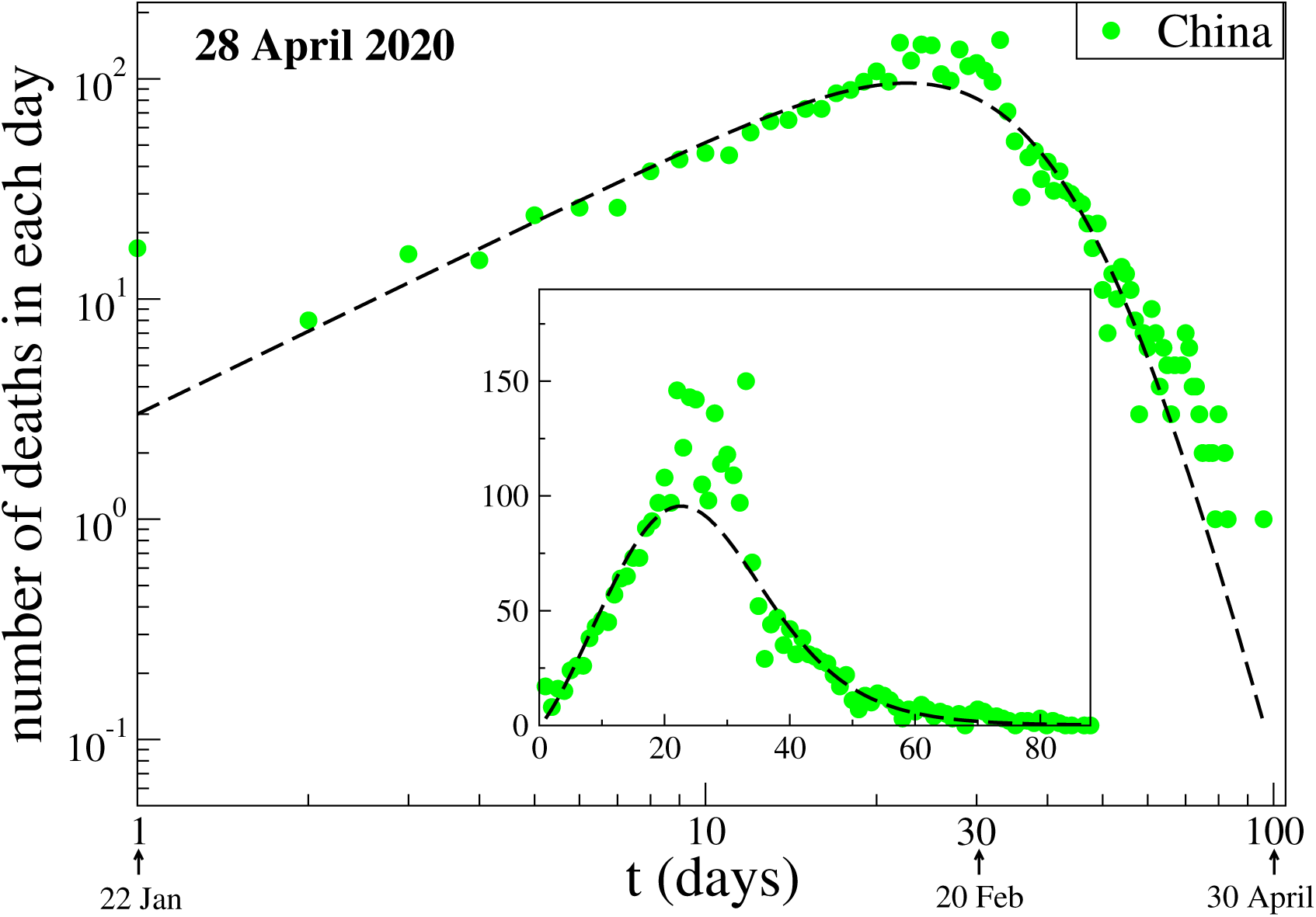
Evolution of deaths per day due to COVID-19 in China. The data are those available at 28 April 2020. The fitting parameters *γ* and *q* are fixed at one of the two choices of the values for China, namely (*γ*, *q*) = (3, 1.26). The other fitting parameters, namely (*C_death_*, *α_death_*, *β_death_*), have been chosen to better fit the available data for China.

**FIG. 4:**
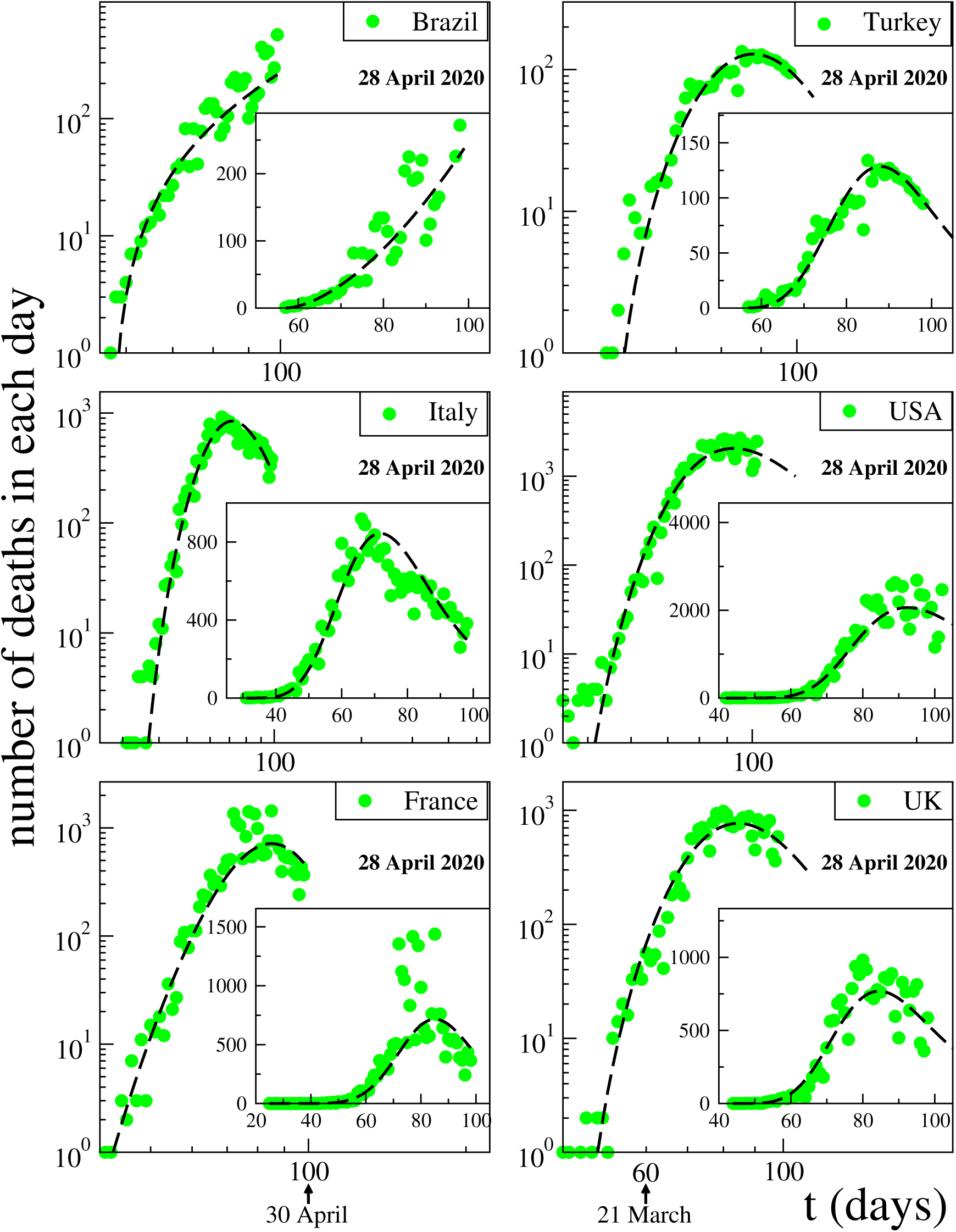
Fittings of deaths per day data for various strongly a ected countries around the world with Eq. (1). The data are those available at 28 April 2020. The fitting parameters *γ* and *q* are fixed at one of the two choices of the values for China, namely (*γ*, *q*) = (3, 1.26). The other fitting parameters, namely (*C*_death_, *α*_death_, *β*_death_), have been chosen to better fit the available data for each country

We may summarise as follows. The death curve of South Korea is atypical, in the sense that it sensibly differs from all those that we analysed in the present work. Because of that, we have not included it here. We remind that the values for (*γ*, *q*) used for the South Korea evolution curve of active cases also differ from those used for all the other countries, which reinforces the fact that some sort of exceptionality exists there, for reasons that are unknown to us. For all countries that do not reach their peak values yet, we have adopted the values of (*γ*, *q*) obtained from the inspection of the entire curve of China.

We verify that the present work appears to belong to the realm of complex systems which includes not only, as mentioned above, high-frequency financial transactions (with *α* > 0 [4, 5], but also anthropological issues such as medieval trading networks and biotech intercorporate networks (with *α* < 0) [14], relaxation in spin-glasses [15], as well as q-Weibull distribution-like systems [16–19], which correspond to the particular case = −1. It remains open as a highly desirable goal to formulate a model which, along lines somewhat similar to epidemiological models such as the SIR one, would predict an evolution curve such as the present Eq. (1). This would clarify important issues such as the conditions under which the values of (*γ*, *q*) could indeed be universal and essentially determined by the biology of the infecting agent, such as the present COVID-19.

## Data Availability

Publicly available datasets were analyzed in this study.

## Acknowledgments

We have benefitted from useful remarks by E.P. Borges and partial support by CNPq and Faperj (Brazilian Agencies).

